# Large-scale empirical identification of candidate comparators for pharmacoepidemiological studies

**DOI:** 10.1101/2023.02.14.23285755

**Authors:** Justin Bohn, James P. Gilbert, Christopher Knoll, David M. Kern, Patrick B. Ryan

## Abstract

**Objectives:** The new user cohort design has emerged as a best practice for the estimation of drug effects from observational data. However, despite its advantages, this design requires the selection and evaluation of comparators for appropriateness, a process which can be challenging. In this paper, we introduce an empirical approach to rank candidate comparators in terms of their similarity to a target drug in high-dimensional covariate space.

**Methods:** We generated new user cohorts for each RxNorm ingredient in five administrative claims databases, then extracted aggregated pre-treatment covariate data for each cohort across five clinically oriented domains. We formed all pairs of cohorts with ≥ 1,000 patients and computed a scalar similarity score, defined as the average of cosine similarities computed within each domain, for each pair. Ranked lists of candidate comparators were then generated for each cohort.

**Results:** Across up to 1,350 cohorts forming 922,761 comparisons, drugs that were more similar in the ATC hierarchy had higher cohort similarity scores. The most similar candidate comparators for each of six example drugs corresponded to alternative treatments used in the target drug’s indication(s), and choosing the top-ranked comparator for randomly selected drugs tended to produce balance on a majority of covariates. This approach also ranked highly those comparators chosen in high quality published new user cohort design studies.

**Conclusion:** Empirical comparator recommendations may serve as a useful aid to investigators and could ultimately enable the automated generation of new user cohort design-derived evidence, a process that has previously been limited to self-controlled designs.

**KEY POINTS:** Empirical comparator recommendations based on similarity to a target cohort’s covariate profile can aid investigators in study design and align with subject matter knowledge and published literature.

## 1 INTRODUCTION

Pharmacoepidemiologic studies routinely employ cohort designs to estimate the effect of a drug treatment on an outcome using observational data. While many variations on the cohort design exist, the new user (NU, sometimes called *incident* new user or *initiator*) design[1,2]has emerged over the past two decades as a best practice. This design, which is intended to emulate a randomized trial in its well-defined, exposure-based timescale, involves identifying cohorts of patients at the time of initiation of one of two drugs: a target drug and a comparator. Patients are then followed for the occurrence of the study outcome and the incidence is compared between groups, usually with adjustment for confounding factors. The NU design can be further divided based on whether an *active* comparator (i.e., an alternative treatment for the indication of interest) or an *inactive* comparator (i.e., something not used for the indication of interest and which is believed to be unrelated to the outcome of interest) is chosen[3]. The primary advantage to this approach is the mitigation of biases known to impact comparisons against nonuser – i.e., unexposed or untreated – groups. In particular, healthy user bias, healthy adherer bias, and immortal time bias may all be reduced by the adoption of an NU design, with particular reductions in confounding by indication expected if an active comparator is chosen[4,5]

However, while the NU design has many advantages, it requires the study designer to select an appropriate comparator, and the process of identifying and determining comparator adequacy can be challenging. The ideal comparator group represents a counterfactual outcome experience determined by the research question of interest. This question may concern whether the incidence of an outcome in the presence of the target exposure is different than what would have occurred if some other exposure were substituted for the target, or alternatively whether that incidence differs from what would have occurred if the target exposure were simply removed. In the former case, which is termed comparative effectiveness or safety research, investigators have historically selected comparators on the basis of perceived clinical equipoise and empirical feasibility. In the latter case, which is common in the drug safety surveillance context, the use of a comparator otherwise thought to be wholly unrelated to the study outcome has been recommended[6–9], and a theoretical framework for its choice developed[10]. While the counterfactual represented by the comparator thus varies according to the research context, a common goal is to identify a comparator with a similar patient composition in terms of demographics and relevant medical characteristics to the target. While several sets of recommendations[11] have emerged to guide investigators in the comparator selection process, it remains at least partially subjective and difficult to scale in a context with many target drugs. With thousands of drugs available on the market, each of which could theoretically be a viable comparator for another, the selection of comparators for all drugs is thus a large task, encompassing nearly one million target-comparator possibilities.

In this paper, we present an empirical approach for ranking potential comparator cohorts for pharmacoepidemiologic studies based on their similarity in high-dimensional covariate space. We propose a method to compute a scalar similarity score, the *cohort similarity score*, for all pairwise combinations of all active drug ingredients and rank candidate comparators for any given target drug based on this composite score. The cohort similarity score is based on the average of cosine similarity scores[12] computed within five separate covariate domains intended to emulate clinical thinking surrounding a treatment decision. These domains are demographics, medical history, prior medications, presentation, and visit context. We evaluate rankings based on the cohort similarity score in the context of existing drug classification schemes and assess whether they produce pairs of cohorts comprised of similar patient populations, based on having high proportions of covariates already in balance.

Throughout, we focus on six example drugs from three therapeutic areas: anticoagulation (warfarin and apixaban), rheumatoid arthritis (methotrexate and adalimumab), and multiple sclerosis (glatiramer and ocrelizumab). We demonstrate that this method produces ranked lists of comparators which broadly align with subject-matter knowledge, and which are already posited as good comparators in the literature or registered studies. Further, we propose that this approach may be of use both as a tool for researchers designing pharmacoepidemiologic studies and as a part of an eventual analytic pipeline to automate the evidence generation process. We provide the full suite of results in an interactive web application (data.ohdsi.org/ComparatorSelectionExplorer), which can be used to identify and diagnose potential comparators for a given target cohort, and the code necessary for others to implement our approach in their own data (https://github.com/OHDSI/ComparatorSelectionExplorer).

## 2 METHODS

### 2.1 Data Sources

We utilized data from five sources of insurance claims: Merative™ MarketScan® Commercial Claims and Encounters (CCAE), Merative™ MarketScan® Multi-State Medicaid (MDCD), Merative™ MarketScan®Medicare Supplemental (MDCR); Japan Medical Data Center (JMDC); and Optum© De-Identified Clinformatics© Data Mart. All data were mapped to the Observational Medical Outcomes Partnership Common Data Model[13] (OMOP CDM). Episodes of continuous exposure to individual RxNorm drug ingredients were inferred from pharmacy claims[14]. International Classification of Diseases (ICD) diagnosis codes were mapped to the Systematized Nomenclature of Medicine Clinical Terms (SNOMED-CT).

### 2.2 Cohort Definitions

Prior to computing similarity scores, it was necessary to create cohorts for each active drug ingredient to be evaluated as either a target or comparator. As such, in each data source, we defined cohorts of new users for each RxNorm ingredient. While there are over 10,000 ingredient concepts in the RxNorm vocabulary, only a minority are used in medical practice and the number varies according to local usage patterns in each data source. To enter a given ingredient-specific cohort, patients were required to have 365 days of continuous observation in the database prior to the first observed exposure to that ingredient, which served as their index date. Patients were allowed to enter a given cohort only once, but there was no restriction on the number of distinct cohorts they could enter. In order to ensure robust estimation of covariate prevalence, we restricted analysis to cohorts with at least 1,000 patients.

### 2.3 Extraction of Covariate Data

Within each cohort, covariate data was extracted in five domains intended to capture the clinical thinking surrounding a treatment decision. These domains were:

- **Demographics**: one covariate for female sex, one for male sex, and one covariate for each decade of age
- **Medical history**: one covariate for each SNOMED condition observed in the period from the start of patient observation to 31 days before the index date
- **Presentation**: one covariate for each SNOMED condition observed in the period from 30 days before the index date to the index date
- **Prior medications:** one covariate for each RxNorm ingredient with exposure start at least 30 days prior to the index date and exposure end on or before the index date
- **Visit context**: one covariate for the presence of an inpatient visit in the 30 days prior to the index date, and one for the presence of an emergency department visit in the same period

Information on the prevalence of each covariate in each cohort was stored in a separate table in the database from which it was extracted and used for the computation of similarity metrics. All covariates with prevalence < 1% were excluded to avoid distortion of similarity scores by extremely low prevalence estimates.

### 2.4 Computation of Similarity Scores

Within each database, we formed all possible pairs of cohorts and computed similarity scores for each pair. We calculated the cosine similarity[12], the dot product of two vectors containing the target and comparator cohorts’ covariate prevalences divided by the product of their lengths, between the two cohorts, separately for each covariate domain, and then averaged across the five domains to get the final similarity score, hereafter called the *cohort similarity score*. While other measures of multivariable similarity/distance such as the average absolute standardized difference and the Mahalanobis distance were available, the computational efficiency of the cosine similarity made it an attractive choice for our large-scale evaluation. A cohort similarity score of 1 will occur if all covariates are the same between two cohorts, while a score of 0 indicates that the cohorts have no agreement across the covariates. We then used these scores to generate ranked lists of comparators for each cohort, with scores closer to 1 indicating stronger comparator alignment.

Under the assumption that cohorts with known clinical or mechanistic relationships should have higher similarity than those without such relationships, we used the Anatomical Therapeutic Chemical (ATC) classification system[15] to compare the distribution of cohort similarity scores for cohort pairs with relationships at different levels of the hierarchy. Cohort pairs were divided according to the most granular ATC level at which they co-occurred, if any. Pairs that co-occurred in a class at ATC level 5 were excluded from this analysis as this level is commonly used for individual drug products or combinations.

For a random subset of 100 cohorts within each data source, we also attempted to validate our ranked lists with a more familiar pharmacoepidemiological metric of covariate similarity, the standardized mean difference (SMD)[16]. For each of the randomly selected target drugs, we identified their top comparator based on the cohort similarity score, calculated SMDs for all covariates present within the cohort pair and reported the proportion of covariates in various degrees of imbalance based on recommended cutoffs for absolute SMD of 0.1, 0.2, 0.5, and 0.8 [17,18]. We then repeated this process for the comparator ranked 500^th^. All code for cohort generation, covariate extraction, similarity calculation, and for our web application is publicly available at https://github.com/OHDSI/ComparatorSelectionExplorer.

### 2.5 Review of NU Design Studies Published in High-Quality Journals

We also searched the literature to identify high-quality observational studies employing NU designs and compared their comparator choices to our empirical estimates of similarity. We limited our search to papers published in the following journals between January 2021 and May 2023: *Science, Nature, The New England Journal of Medicine*, *JAMA, JAMA Internal Medicine, JAMA Surgery, JAMA Psychiatry, JAMA Cardiology, JAMA Pediatrics, JAMA Neurology, The Lancet, BMJ, Diabetes Care, Hypertension, Journal of the American College of Cardiology, Pharmacoepidemiology and Drug Safety, Drug Safety, Annals of Internal Medicine, The Lancet Digital Health, The Lancet Infectious diseases, The Lancet Diabetes & Endocrinology, The Lancet Neurology, The Lancet Oncology, The Lancet Respiratory Medicine, The Lancet Psychiatry, and The Lancet Global Health*. Full details on the search parameters used and the results of the review can be found in the supplementary material. As some of the identified studies defined target/comparator cohorts at the drug *class* level, we additionally generated new-user cohorts for each Anatomical Therapeutic Chemical (ATC) level 4 class and re-ran our similarity pipeline with these cohorts included.

## 3 RESULTS

Between 1,729 and 2,409 RxNorm ingredient-level new-user cohorts were generated per data source. Of these, between 988 and 1,359 cohorts had at least 1,000 patients (Table 1). The mean number of covariates with prevalence ≥1% extracted per cohort was 934.5 - 1,479.9 overall, 65.4-120.0 in the “Presentation” domain, 336.9-1,016.4 in the “Medical History” domain, and 293.0 - 486.2 in the “Prior Medications” domain. The number of demographic and visit context covariates was roughly constant by design.

**Table 1:** Summary of cohorts generated, and covariates extracted, by data source and covariate domain.

|  | CCAE | MDCD | MDCR | JMDC | Optum |
| --- | --- | --- | --- | --- | --- |
| **Total number of cohorts generated** |  |  |  |  |  |
| *Overall* | 2,243 | 2,158 | 2,065 | 1,729 | 2,273 |
| *With at least 1,000 patients* | 1,359 | 1,112 | 988 | 1,082 | 1,332 |
| **Mean (SD) number of covariates with prevalence $\geq 1\%$ extracted per cohort with $\geq 1,000$ patients** |  |  |  |  |  |
| *All* | 1,030.6 (324.65) | 1,479.9 (432.59) | 1,287.7 (279.28) | 934.5 (186.76) | 1,422.0 (441.61) |
| *Demographics* | 7.8 (1.12) | 9.6 (1.52) | 6.2 (0.57) | 8.9 (1.23) | 9.3 (1.64) |
| *Medical history* | 662.3 (234.73) | 1,016.4 (319.43) | 852.1 (207.44) | 336.9 (74.25) | 957.3 (309.51) |
| *Presentation* | 65.4 (49.17) | 120.0 (63.71) | 88.3 (47.32) | 101.5 (36.33) | 119.2 (85.42) |
| *Prior medications* | 293.0 (58.56) | 331.9 (67.89) | 339.1 (39.66) | 486.2 (83.36) | 334.3 (64.94) |
| *Visit context* | 1.8 (0.36) | 1.9 (0.25) | 1.9 (0.08) | 1.0 (0.00) | 1.8 (0.34) |
| **Number of pairs of cohorts with $\geq 1,000$ patients** | 922,761 | 617,716 | 487,578 | 584,821 | 886,446 |
| **Mean (SD) similarity score among pairs of cohorts with $\geq 1,000$ patients** |  |  |  |  |  |
| *Cohort similarity score* | 0.721 (0.099) | 0.774 (0.106) | 0.847 (0.060) | 0.793 (0.090) | 0.743 (0.120) |
| *Demographics cosine similarity* | 0.852 (0.139) | 0.846 (0.128) | 0.945 (0.075) | 0.857 (0.134) | 0.841 (0.147) |
| *Medical history cosine similarity* | 0.781 (0.137) | 0.798 (0.137) | 0.913 (0.059) | 0.879 (0.092) | 0.798 (0.141) |
| *Presentation cosine similarity* | 0.213 (0.173) | 0.401 (0.216) | 0.490 (0.190) | 0.377 (0.189) | 0.349 (0.212) |
| *Prior medications cosine similarity* | 0.882 (0.085) | 0.872 (0.090) | 0.931 (0.059) | 0.917 (0.071) | 0.859 (0.108) |
| *Visit context cosine similarity* | 0.875 (0.150) | 0.955 (0.066) | 0.957 (0.069) | 1.000 (0.000) | 0.869 (0.160) |
| **Number (%) of target cohorts with at least one comparator with cohort similarity score of:** |  |  |  |  |  |
| *At least 0.900* | 1,294 (95.2%) | 1,094 (98.4%) | 981 (99.3%) | 1,067 (98.6%) | 1,294 (97.1%) |
| *At least 0.950* | 1,087 (80.0%) | 961 (86.4%) | 924 (93.5%) | 1,024 (94.6%) | 1,118 (83.9%) |

CCAE: Merative™ MarketScan® Commercial Claims and Encounters; MDCD: Merative™ MarketScan® Multi-State Medicaid; MDCR: Merative™ MarketScan® Medicare Supplemental; JMDC: Japan Medical Data Center; Optum: Optum© De-Identified Clinformatics© Data Mart. Cohort similarity score is average of domain-specific cosine similarities.

Between 487,578 and 922,761 pairs of cohorts with at least 1,000 patients were formed per data source. Among these, similarity was lowest in the “Presentation” domain (mean cosine similarity 0.213 to 0.490, depending on data source) and highest in the “Visit context” domain (mean cosine similarity 0.869 to 1.000, depending on data source). Between 90% and 95% of target cohorts with at least 1,000 patients had a comparator with an average cohort similarity score of ≥ 0.950. Among the six example drugs, the distribution of cohort similarity scores was consistently left-skewed, with most comparators lying between 0.7 and 0.9 (Figure 1). Similarity was lowest among pairs of cohorts that did not occur together in any ATC classes and tended to increase among pairs of cohorts that co-occurred within higher-level ATC classes (Figure 2).

**Figure 1:**
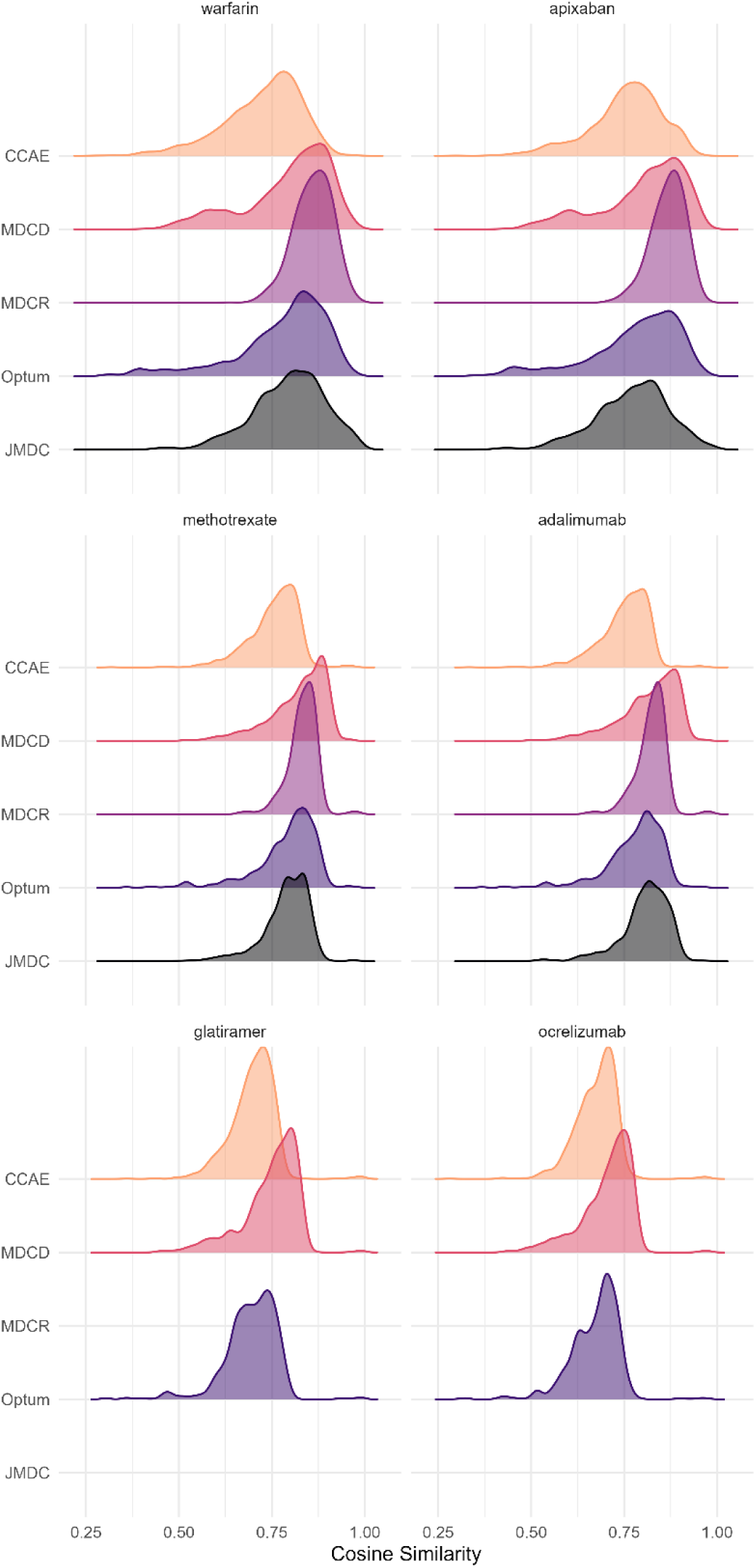
Distribution of average cosine similarity scores computed across all available candidate comparators for six example drugs, by data source. Note that glatiramer and ocrelizumab did not meet the 1,000 patient sample size cutoff in MDCR or JMDC and thus results are not present for these data sources. The number of available comparators ranged from 988-1,359, depending on data source.

**Figure 2:**
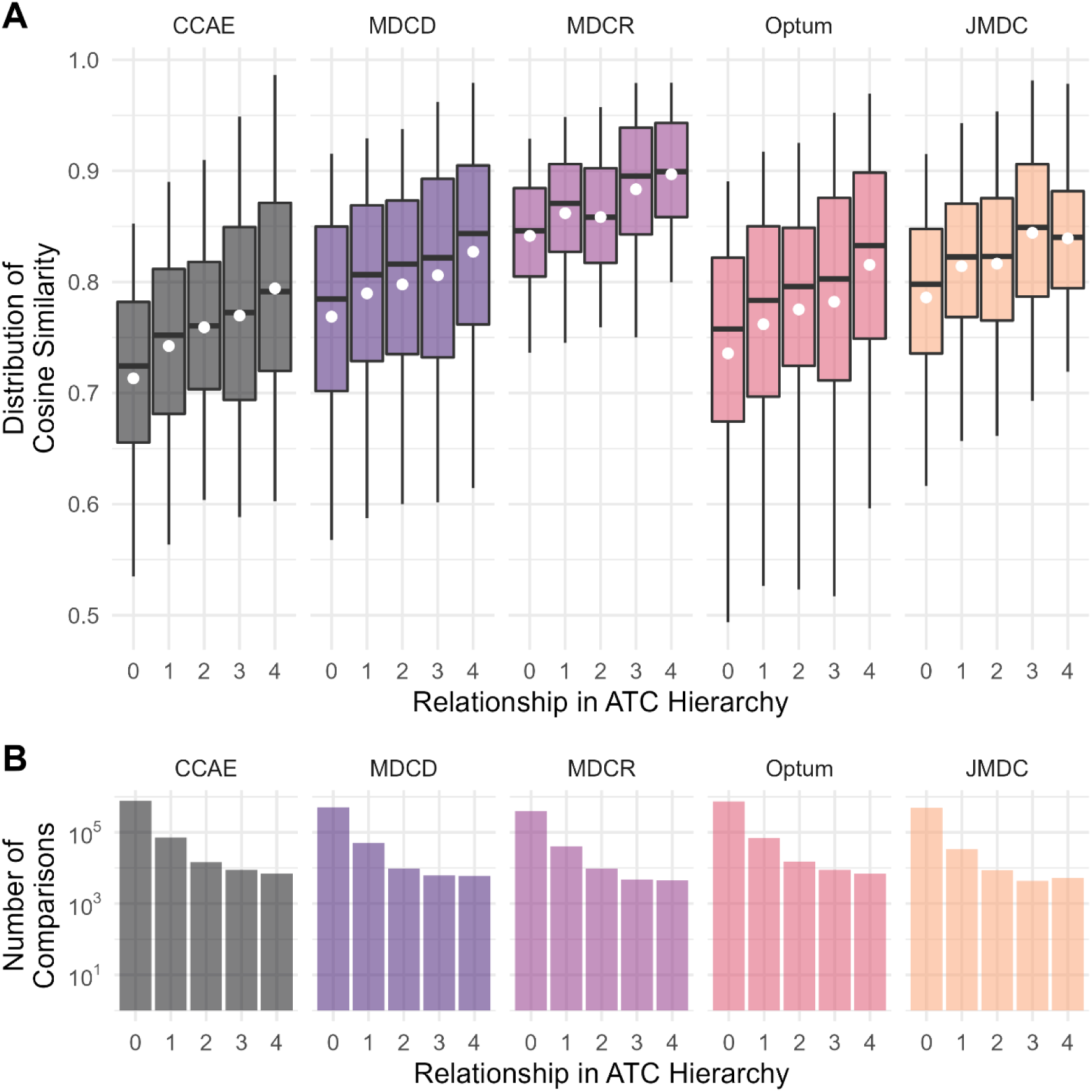
Distribution of cohort similarity scores (panel A) and number of comparisons (panel B), by relatedness in ATC hierarchy and data source. All target-comparator pairs were stratified by the distance between the two drugs in the ATC hierarchy (“4”, indicating that both drugs are in same low-level class, through “0”, indicating that the drugs do not share any common drug class ancestry). Cohort pairs in which the target and comparator are both members of an ATC level 5 class are omitted as these classes are frequently used for individual drugs or combination products. Whiskers are truncated at 10^th^ and 90^th^ percentiles. White circles indicate mean values.

The top five comparators for each of the six example drugs, by average rank across data sources (Table 2) and within data sources (Table 3) consistently corresponded to alternative treatments for each drug’s indication(s). For example, the top five comparators for glatiramer (interferon beta-1a, interferon beta-1b, dimethyl fumarate, fingolimod, and natalizumab) are all other disease-modifying therapies for multiple sclerosis[19], and those ranked sixth through tenth (teriflunomide, dalfampridine, ocrelizumab, corticotropin, and modafinil) are either disease modifying therapies or are used to treat symptoms of MS or MS relapse. Ranked comparator lists for these example drugs varied according to data source (Figure 1, Figure 3 panel A), with the highest degree of correlation between CCAE, MDCD, and Optum. An example of between-data source heterogeneity is shown in Figure 3 panel B where, among 627 candidate comparators for warfarin observed in both CCAE and JMDC, rivaroxaban is identified as highly similar to warfarin in both data sources, whereas xylitol and vitamin A appear much more similar to warfarin in JMDC than in CCAE, while the reverse is true for hydralazine.

**Figure 3:**
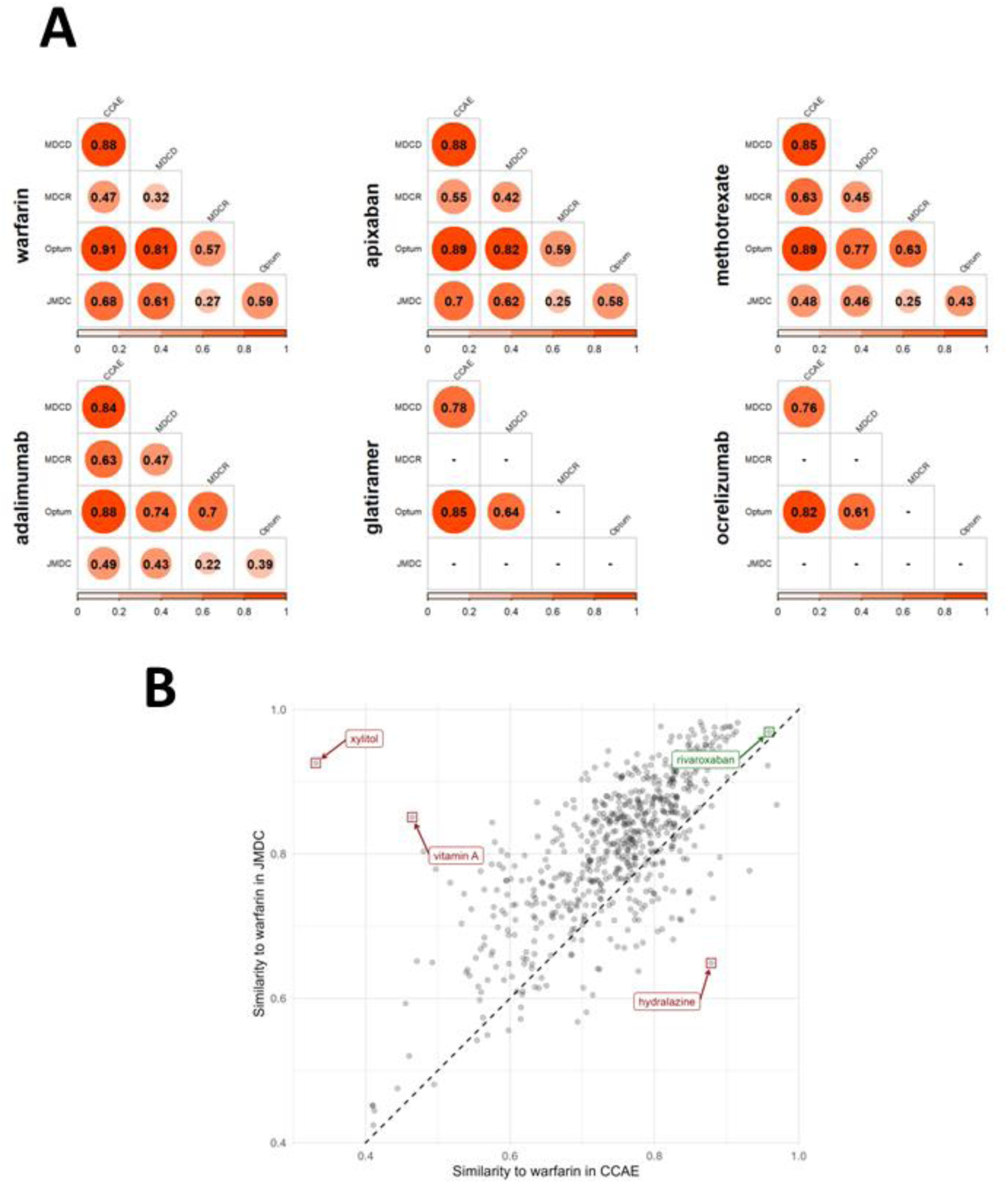
Pairwise rank correlations in cohort similarity scores between data sources for six example drugs (A) and cosine similarity scores among 627 candidate comparators for warfarin observd in both CCAE and JMDC (B).

**Table 2:** Top five comparators for six example drugs, according to average rank across five data sources.

| Rank | warfarin | apixaban | methotrexate | adalimumab | glatiramer | ocrelizumab |
| --- | --- | --- | --- | --- | --- | --- |
| 1 | digoxin (4.6, 0.960, 5) | rivaroxaban (1.0, 0.977, 5) | sulfasalazine (1.2, 0.978, 5) | certolizumab pegol (2.2, 0.974, 4) | interferon beta-1a (1.3, 0.993, 3) | teriflunomide (1.0, 0.980, 2) |
| 2 | rivaroxaban (6.2, 0.968, 5) | diltiazem (7.0, 0.955, 5) | leflunomide (2.5, 0.970, 4) | sulfasalazine (3.4, 0.960, 5) | interferon beta-1b (1.5, 0.993, 2) | dimethyl fumarate (2.3, 0.969, 3) |
| 3 | diltiazem (6.4, 0.959, 5) | amiodarone (9.6, 0.953, 5) | etanercept (3.4, 0.966, 5) | infliximab (4.0, 0.958, 5) | dimethyl fumarate (3.3, 0.991, 3) | natalizumab (3.0, 0.967, 3) |
| 4 | carvedilol (9.0, 0.952, 5) | flecainide (14.2, 0.952, 5) | adalimumab (4.6, 0.953, 5) | methotrexate (4.2, 0.953, 5) | fingolimod (3.3, 0.989, 3) | fingolimod (3.7, 0.965, 3) |
| 5 | amiodarone (10.2, 0.953, 5) | torsemide (14.4, 0.947, 5) | certolizumab pegol (6.5, 0.953, 4) | leflunomide (6.0, 0.945, 4) | natalizumab (4.3, 0.988, 3) | dalfampridine (4.0, 0.967, 2) |

2 Restricted to comparators occurring in at least two data sources. Entries are given as comparator name (average rank across data sources, mean 3 cohort similarity score, number of data sources with information on comparison).

**Table 3:** Top five comparators for six example drugs according to cohort similarity score, by data source.

| Example Drug | Data Source | Comparator #1 | Comparator #2 | Comparator #3 | Comparator #4 | Comparator #5 |
| --- | --- | --- | --- | --- | --- | --- |
| warfarin | CCAE | fondaparinux (0.970) | rivaroxaban (0.959) | dalteparin (0.957) | enoxaparin (0.932) | digoxin (0.915) |
| warfarin | MDCD | diltiazem (0.973) | carvedilol (0.970) | dabigatran etexilate (0.970) | rivaroxaban (0.968) | captopril (0.968) |
| warfarin | MDCR | diltiazem (0.975) | digoxin (0.972) | rivaroxaban (0.971) | amiodarone (0.971) | sotalol (0.970) |
| warfarin | Optum | rivaroxaban (0.970) | dabigatran etexilate (0.967) | captopril (0.967) | diltiazem (0.966) | digoxin (0.965) |
| warfarin | JMDC | protamines (0.988) | dexmedetomidine (0.986) | norepinephrine (0.985) | dobutamine (0.983) | digoxin (0.982) |
| apixaban | CCAE | rivaroxaban (0.956) | protamine sulfate (USP) (0.951) | digoxin antibodies<br>Fab fragments (0.940) | diltiazem (0.940) | hydralazine (0.935) |
| apixaban | MDCD | rivaroxaban (0.985) | torsemide (0.967) | diltiazem (0.967) | bumetanide (0.966) | carvedilol (0.964) |
| apixaban | MDCR | rivaroxaban (0.974) | diltiazem (0.952) | hydralazine (0.952) | enoxaparin (0.952) | protamine sulfate (USP) (0.951) |
| apixaban | Optum | rivaroxaban (0.973) | digoxin antibodies<br>Fab fragments (0.972) | amiodarone (0.971) | diltiazem (0.968) | torsemide (0.966) |
| apixaban | JMDC | rivaroxaban (0.998) | dabigatran etexilate (0.990) | digoxin (0.988) | edoxaban (0.985) | flecainide (0.985) |
| methotrexate | CCAE | sulfasalazine (0.972) | leflunomide (0.970) | etanercept (0.969) | hydroxychloroquine (0.956) | adalimumab (0.955) |
| methotrexate | MDCD | sulfasalazine (0.961) | etanercept (0.960) | hydroxychloroquine (0.955) | adalimumab (0.954) | leflunomide (0.950) |
| methotrexate | MDCR | leflunomide (0.984) | sulfasalazine (0.984) | hydroxychloroquine (0.981) | adalimumab (0.980) | etanercept (0.979) |
| methotrexate | Optum | sulfasalazine (0.983) | leflunomide (0.976) | hydroxychloroquine (0.974) | etanercept (0.958) | adalimumab (0.955) |
| methotrexate | JMDC | sulfasalazine (0.988) | tocilizumab (0.968) | etanercept (0.966) | golimumab (0.962) | adalimumab (0.922) |
| adalimumab | CCAE | certolizumab pegol (0.973) | etanercept (0.962) | sulfasalazine (0.957) | methotrexate (0.955) | infliximab (0.948) |
| adalimumab | MDCD | sulfasalazine (0.970) | certolizumab pegol (0.970) | etanercept (0.960) | methotrexate (0.954) | infliximab (0.942) |
| adalimumab | MDCR | etanercept (0.990) | leflunomide (0.983) | infliximab (0.982) | methotrexate (0.980) | certolizumab pegol (0.978) |
| adalimumab | Optum | certolizumab pegol (0.973) | etanercept (0.960) | sulfasalazine (0.959) | methotrexate (0.955) | golimumab (0.948) |
| adalimumab | JMDC | infliximab (0.982) | azathioprine (0.963) | golimumab (0.956) | sulfasalazine (0.939) | methotrexate (0.922) |
| glatiramer | CCAE | interferon beta-1a (0.994) | interferon beta-1b (0.993) | fingolimod (0.992) | natalizumab (0.992) | dimethyl fumarate (0.991) |
| glatiramer | MDCD | interferon beta-1a (0.994) | dimethyl fumarate (0.992) | fingolimod (0.987) | natalizumab (0.986) | ocrelizumab (0.962) |
| glatiramer | Optum | interferon beta-1b (0.992) | interferon beta-1a (0.991) | dimethyl fumarate (0.989) | fingolimod (0.989) | natalizumab (0.987) |
| ocrelizumab | CCAE | teriflunomide (0.981) | dimethyl fumarate (0.974) | fingolimod (0.969) | natalizumab (0.968) | peginterferon beta-1a (0.965) |
| ocrelizumab | MDCD | natalizumab (0.977) | dimethyl fumarate (0.975) | fingolimod (0.974) | glatiramer (0.962) | interferon beta-1a (0.953) |
| ocrelizumab | Optum | teriflunomide (0.979) | dalfampridine (0.971) | dimethyl fumarate (0.960) | natalizumab (0.957) | fingolimod (0.952) |

1 Entries are given as comparator name (cohort similarity score). Note that glatiramer and ocrelizumab did not meet the 1,000 patient sample size 2 cutoff in MDCR or JMDC and thus results are not present for these data sources.

Among the 100 target drugs randomly selected within each data source and their top comparators, most covariates were well balanced. When the comparator ranked 500^th^ was chosen instead, the proportion of imbalanced covariates was higher (Figure 4). The average proportion of covariates with |SMD| ≤ 0.1 was between 72% and 84% for the top comparator versus between 28% and 45% for the comparator ranked 500^th^.

**Figure 4:**
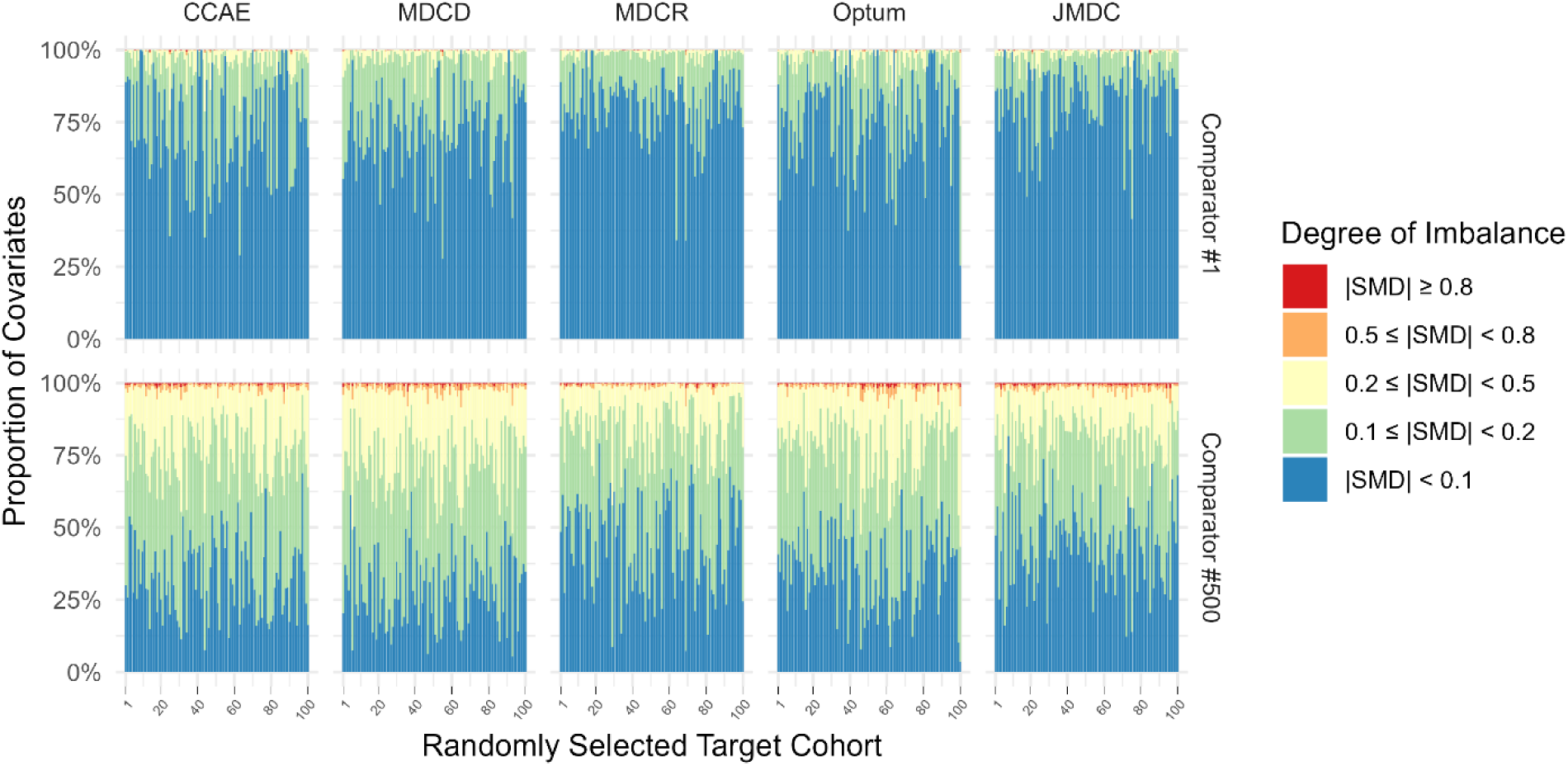
Proportion of covariates at various levels of imbalance according to standardized mean difference (SMD) among 100 randomly-selected target cohorts and their comparators ranked 1^st^ and 500^th^, by data source.

Our literature search returned 60 studies, of which 24 employed an NU design. 56 distinct target-comparator pairs were identified, of which 23 could be mapped directly to RxNorm ingredients or ATC level 4 classes. Nine such pairs were comparisons among anticoagulants, and five were among antidepressants. Among these 20 pairs, the comparator selected by the authors was in the top 5% of computed similarity for its target in at least one of our data sources in 96% of cases, and in 91% of cases it was ranked among the top 10. Some pairs with notably low computed similarity were apixaban vs. warfarin (cohort similarity score 0.879, rank 136 in CCAE), beta blocking agents vs. aldosterone antagonists (cohort similarity score 0.757, rank 359 in CCAE), and topiramate vs. lamotrigine (cohort similarity score 0.846, rank 167 in CCAE). See supplementary file for similarity scores from reviewed literature.

## 4 DISCUSSION

Over the past two decades, the NU cohort design has emerged as a best practice for the estimation of drug effects from observational data. However, the implementation of this design may be hampered by the difficulty of selecting an appropriate comparator for the target drug of interest, either because there are many potential alternatives to the target drug, or because the true target of inference is in fact a contrast between the target drug and its absence. The development of empirical comparator recommendations can aid investigator decision-making, simplifying the process of comparator selection and improving the quality of pharmacoepidemiological research and the speed of postmarketing surveillance. Ultimately, these approaches may even enable the automation of evidence generation using the NU design, something that was previously possible only with self-controlled designs[20,21] and temporal scans[22,23].

We demonstrated a method for ranking potential comparators based on similarity in high-dimensional covariate space. We showed that most target cohorts of reasonable size possessed at least one potential comparator with a high degree of similarity and that cohorts that were more related in the ATC hierarchy also had higher cohort similarity scores. For a random sample of 100 cohorts within each data source, we also showed that selecting the top-most similar comparator identified a cohort with balance on the majority of covariates before any adjustment took place.

We additionally demonstrated that, for six target cohorts from a diverse set of therapeutic areas, the top-ranked comparators aligned well with subject-matter knowledge. That is, all were used in the treatment of at least one of the target cohorts’ indications. Indeed, many of these top-ranked comparators are found to be in use in head-to-head safety or effectiveness comparisons with the target cohort in the literature or in public registries of clinical and observational studies. For example, head-to-head studies exist comparing apixaban and its top comparator rivaroxaban[24,25], adalimumab and its top comparator certolizumab[26,27], and ocrelizumab and its top comparator teriflunomide[28]. This was also borne out in our formal literature review, which demonstrated that NU studies published in high quality journals tended to select comparators that our method also ranked highly. While this is reassuring, it should also be noted that the correlation in comparator rankings across data sources was not consistently high (Figure 3). This may indicate that a given comparator is not uniquely fit-for-purpose across all data sources, suggesting that investigators take care to select a comparator using information from a source that at least resembles their intended analytic context.

This study complements a large body of work on drug similarity from the bioinformatics community, including attempts to define similarity based on chemical structure, gene expression[29], genome-wide associations[30], co-occurrence of drugs with condition terms in the literature[31], individual patient data[32], and a combination of these. However, our work is unique in its focus on the use of similarity indices for comparator selection rather than for identification of novel associations, and for its clinically oriented approach to covariate data extraction. The empirical comparator recommendations provided here have several advantages. First, they can reduce subjectivity, both in terms of which drugs may serve as candidate comparators for a given target drug and which covariates should be used to determine empirical feasibility. While comparator choice should be guided by subject-matter knowledge[33], no investigator is omniscient and thus the availability of empirical recommendations may promote more informed decisions and could potentially enable less biased comparisons. Additionally, empirical recommendations may reveal previously unknown comparators that are not necessarily alternative treatments for any of the target drug’s indications. They thus hold promise for the identification of appropriate inactive comparators/negative control exposures, ultimately allowing for cohort studies that emulate user versus non-user comparisons within the NU framework. Additionally, these recommendations have been assessed across multiple large data sources comprising diverse populations within and outside the United States. They have been observed, consistently across these data sources, to label drugs that are more closely related in the ATC hierarchy as more similar, to produce balance on a majority of covariates when the most similar comparator is chosen, and to identify top comparators that align with subject-matter knowledge and high quality published research. Finally, the approach presented here provides a standardized framework for comparator ranking which can be readily implemented by investigators. This standardization can improve reproducibility and scalability and ultimately lowers the burden on domain expertise for the investigator.

In an era of rapid advancement in deep learning methods, the simplicity of our approach may appear to be a limitation. For example, emerging research on large language models (LLMs) [34] may suggest their potential utility as a source of comparator recommendations, and much has been written on the utility of various similarity-based deep learning approaches to predict drug-drug and drug-target interactions. Unfortunately, the lack of a reference corpus explicitly identifying “good” and “bad” comparators prevents us from trialing these methods here. And while we look forward to future assessments of their utility, LLM approaches provide little empirical insight as to the origin of their responses, can suffer from hallucinations, and are naturally biased towards pre-existing information, including non-refereed online context, reducing their utility in identifying negative control exposures or previously unused but otherwise suitable comparators.

Compared to traditional approaches, a limitation of the approach introduced here is that the cohort similarity score is based on the *marginal* distributions of covariates in two cohorts and as such is only a proxy for full covariate similarity. In contrast, propensity scores methods have the advantage of providing metrics (e.g., the c-statistic or the preference score[35]) that reflect the similarity in *joint* covariate distributions across cohorts. Furthermore, such methods can be used to adjust for observed dissimilarity when it occurs. However, unlike many propensity score models, which only incorporate a small number of covariates, the cohort similarity score dynamically defines a large set of covariates based on all available data elements. It should be noted that this data-adaptive approach has been incorporated into some propensity score methods like the high-dimensional propensity score (hdPS) [36] and, more recently, the large-scale propensity score (LSPS)[37]. However, the cohort similarity score remains the only approach that is scalable to millions of candidate target-comparator combinations, whereas PS models—particularly those using high-dimensional covariate data—can be computationally expensive, making them infeasible for screening large numbers of candidate comparators. For example, on an Amazon EC2 instance running Windows Server 2019 with a 2.2 GHz processor and 64 GB RAM and accessing data stored in a Redshift database, our entire pipeline (including generating cohorts, extracting covariate data, and computing cohort similarity scores for all drugs) took between 33 and 172 minutes per database, whereas an LSPS pipeline (including generating cohorts, extracting covariate data, and fitting models) took between 3 and 40 hours *for a single target-comparator pair.* The cohort similarity score may therefore be best suited as a tool to narrow the comparator search space before techniques like hdPS/LSPS are employed. It should also be noted that, like PS methods and assessments of standardized differences in covariates, our approach only addresses half of the confounding picture. That is, it summarizes covariate imbalance without addressing covariates’ associations with study outcomes. And while investigators must keep this limitation in mind, they can address it *post-hoc* by searching our web application to identify the degree of balance on important risk factors for their study outcome of interest.

There are several factors that may limit the usefulness of the comparator recommendations provided here. Because this analysis was limited to cohorts of new users of individual RxNorm ingredients, comparator recommendations for ingredients that are used differently depending on dose, form, or route of administration may be less coherent than recommendations for ingredients which are used more consistently regardless of these factors. The same limitation may apply to ingredients that are used in combination preparations or which possess multiple, possibly unrelated, indications. However, this can be ameliorated by running the entire analytic pipeline in the subset of the population with a specific indication of interest. A demonstration of this for the target spironolactone, which is used both in the treatment of acne and heart failure, showed that such restriction both reduces the comparator search space and improves the coherence of recommendations (see supplementary material for details). While such restriction is not feasible at the scale of all indications, it may be attractive to investigators whose work routinely focuses on a small set of indications. Additionally, the assessment of these recommendations was limited by the lack of an objective metric for defining comparator “appropriateness”. While we sought to benchmark appropriateness based on comparator location in the ATC hierarchy and appearance in published literature or registered studies, future work might attempt to go further by demonstrating that the use of empirical comparator recommendations can lead to less biased treatment effect estimates than traditional methods of comparator selection.

Finally, it should be clearly stated that empirical comparator recommendations are not a replacement for critical thinking in study design and analysis. Investigators may wish to use them as a “first pass” to identify and evaluate candidate comparators, after which more careful consideration and detailed analysis can determine true feasibility. Crucially, investigators must remain aware that covariate adjustment is almost invariably required regardless of the quality of the comparator chosen. Similarity between a target drug and comparator is also no guarantee that any remaining covariate imbalance can be successfully adjusted via established methods like restriction or propensity score adjustment. And, conversely, low similarity relative to other recommendations is not a guarantee that successful adjustment is impossible.

Our work leaves open many exciting opportunities for further exploration of empirical comparator recommendations. Among these are additional approaches to validation, including a comparison against more computationally expensive techniques like assessing propensity score overlap. Future work might also explore the utility of adding sources of metadata on known or suspected drug-outcome relationships, such as the OHDSI Common Evidence Model[38,39], in order to improve the ease of identifying inactive comparators/negative control exposures.

## 5 CONCLUSION

We have developed and evaluated a method for recommending comparators that yields promising results and which may be of use to investigators wishing to conduct NU studies of drug safety and effectiveness. An open-source tool set is available for those wishing to identify candidate comparators using their own data, and the full suite of results, including comparator rankings for thousands of drugs across multiple data sources, is available to the public. We hope these tools and the results presented here will accelerate and improve the quality of future pharmacoepidemiologic research.

## Supporting information

Supplementary material: indication restriction demonstration

Supplementary material: literature review methods

Supplementary material: literature review results

## DECLARATIONS

### Data availability

The full suite of results described here is available as an interactive web application (data.ohdsi.org/ComparatorSelectionExplorer), which can be used to identify and diagnose potential comparators for a given target cohort. Note that this application undergoes periodic updates, including the updating of existing data sources to newer versions, the addition of new data sources, and the modification of existing functionality. As such, some results derived from the application may differ from those presented here.

### Code availability

All code necessary to run the analytic pipeline described here and to generate the web application is available via GitHub (https://github.com/OHDSI/ComparatorSelectionExplorer).

### Author contributions

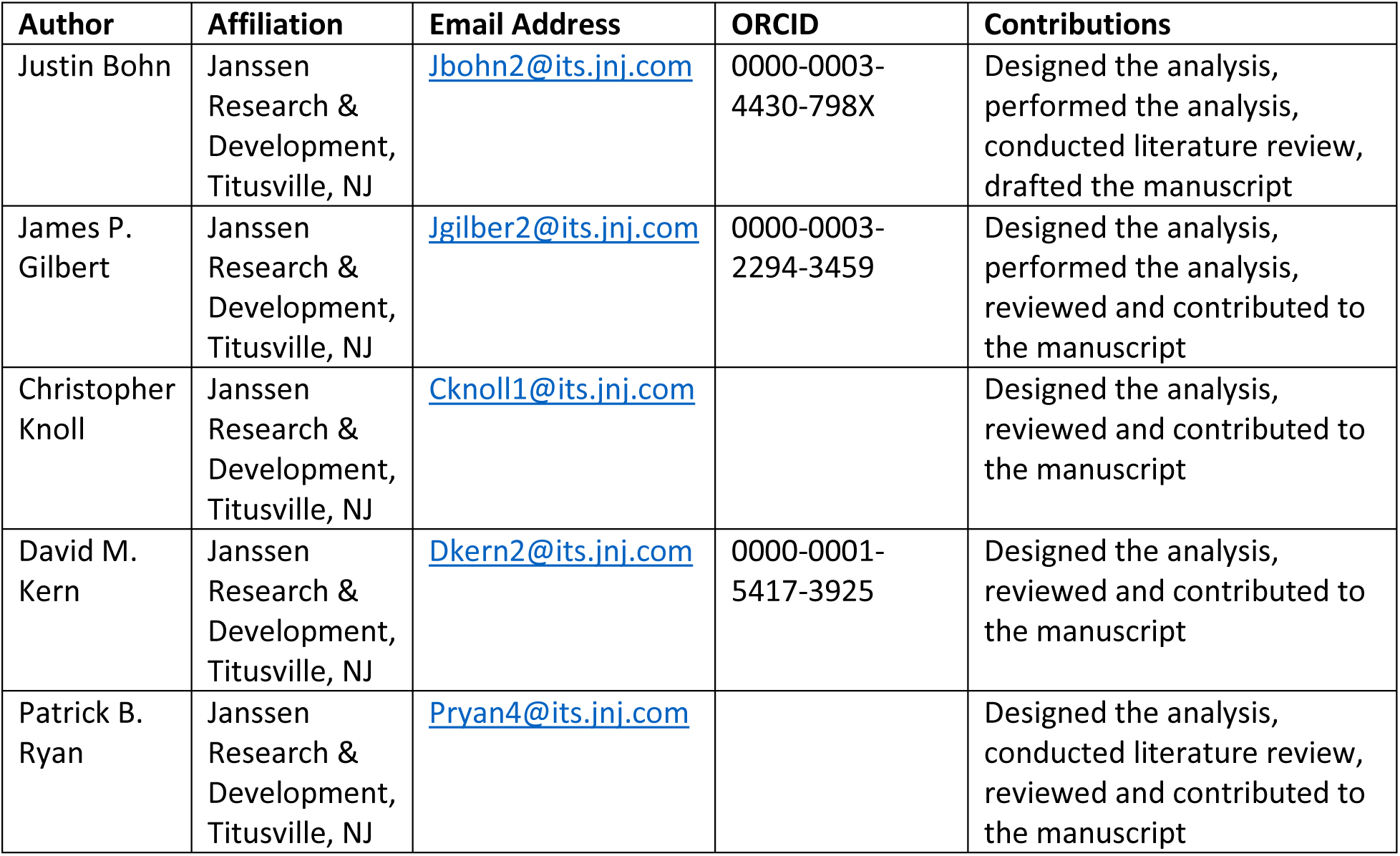

