## Supplementary material: indication restriction demonstration for "Large-scale empirical identification of candidate comparators for pharmacoepidemiological studies"

[1] Global Epidemiology Organization
Janssen Research and Development, LLC, Titusville, NJ, USA

Correspondence to:
Justin Bohn
1125 Trenton-Harbourton Road
Titusville, NJ, USA, 08560

+1 (857) 772-2015

### Supplemental Material: Demonstration of Indication Restriction

#### Description and Methodology

One of the limitations of the recommendations generated by our approach is that their coherence may suffer when the target drug has multiple disparate indications. One potential solution to this problem is to run the entire analytic pipeline separately within sub-populations who meet the criteria for each indication. And while it is not feasible to do this at the scale of all indications, this may be an attractive approach to an investigator focused on one or a few specific areas.

We demonstrate this for the target drug spironolactone, which is used to treat high blood pressure and reduce the risk of mortality in patients with heart failure (HF), but is also used off-label for the treatment of acne. Using data from the Merative™ MarketScan® Commercial Claims and Encounters (CCAE) database, we generated empirical comparator recommendations overall, and separately among patients with diagnosed HF and with diagnosed acne within the 365 days prior to initiation of spironolactone or a comparator drug.

#### Results

Without restriction based on indication there were 905,536 initiators of spironolactone, and 1,476 potential ingredient-based comparators with ≥ 1,000 patients in the CCAE database. After restriction of all cohorts to patients with HF, target sample size was reduced to 97,993 and the number of comparator drugs with ≥ 1,000 patients was reduced to 658. After restriction of all cohorts to patients with acne, target sample size was reduced to 331,062 and the number of comparators with ≥ 1,000 patients was reduced to 723. The distribution of cohort similarity scores among comparators for spironolactone, both with and without indication restriction is shown in **Supplemental figure 1**. The top five comparators ranked according to cohort similarity score, both with and without indication restriction are provided in **Supplemental table 1**.

### Discussion

Because the HF and acne populations differ substantially in their demographic and medical profiles, the list of top-ranked comparators for spironolactone generated without indication restriction does not represent a coherent set of treatments related to either condition. By restricting based on indication, the comparator search space is narrowed and the indication-restricted comparator cohorts that remain exhibit a higher degree of similarity to the target and less inter-comparator variability. This restriction also results in more coherent recommendations. For example, after restriction to the HF population, the top ranked comparators include other HF treatments such as the beta blockers carvedilol and bisoprolol, the ACE inhibitor lisinopril, and the loop diuretics furosemide and torsemide. Likewise, after restriction to the acne population, the top ranked comparators include other acne treatments such as the antibacterials azelate and dapsone and the corticosteroid triamcinolone, as well as the contraceptives levonorgestrel and etonogestrel, which are frequently prescribed with spironolactone. While this indication-restricted approach is not feasible at the scale of all indications, it may have utility for an investigator focused on one or a few specific indications. Alternatively, organizations tasked with the study of several targets nested within a smaller number of indications (e.g., pharmaceutical companies) may benefit from running the similarity pipeline described in the main text separately within each indication under their purview and with restriction to only those comparisons involving the relevant targets of interest, thereby saving computation time and increasing the utility of the resulting empirical recommendations.

### Figures

*Supplemental figure 1: Distribution of cohort similarity scores among comparators for spironolactone, with and without restriction to heart failure and acne populations*

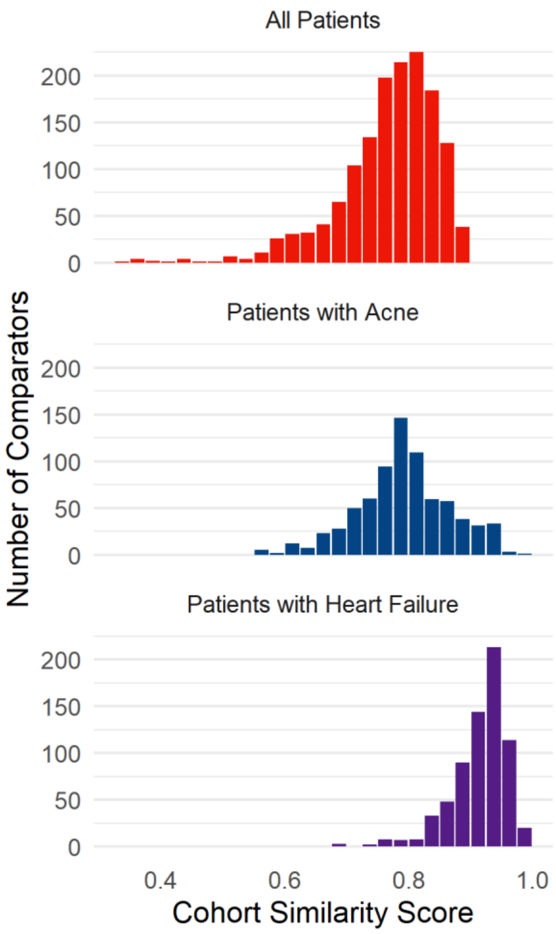

### Tables

*Supplemental table 1: Top-ranked comparators for spironolactone, overall and with restriction according to indication*

| Rank | All Patients | Patients with Heart Failure | Patients with Acne |
|---|---|---|---|
| 1 | furosemide (0.897) | carvedilol (0.992) | azelate (0.978) |
| 2 | clindamycin (0.895) | bisoprolol (0.990) | dapsone (0.969) |
| 3 | Lactobacillus acidophilus (0.895) | torsemide (0.988) | triamcinolone (0.956) |
| 4 | ethacrynate (0.894) | furosemide (0.987) | levonorgestrel (0.952) |
| 5 | enoxaparin (0.892) | lisinopril (0.986) | etonogestrel (0.947) |

Comparators presented as *ingredient name (cohort similarity score)*
