## Supplementary material: literature review methods for "Large-scale empirical identification of candidate comparators for pharmacoepidemiological studies"

[1] Global Epidemiology Organization
Janssen Research and Development, LLC, Titusville, NJ, USA

Correspondence to:
Justin Bohn
1125 Trenton-Harbourton Road
Titusville, NJ, USA, 08560

+1 (857) 772-2015

### Supplemental Material: Review of Published New User Cohort Design Studies

#### Description and Methodology

We searched the literature to identify high-quality observational studies employing new user (NU) designs and compared their comparator choices to our empirical estimates of similarity. We limited our search to papers published in the following journals in 2021 or later: *Science, Nature, The New England Journal of Medicine*, *JAMA, JAMA Internal Medicine, JAMA Surgery, JAMA Psychiatry, JAMA Cardiology, JAMA Pediatrics, JAMA Neurology, The Lancet, BMJ, Diabetes Care, Hypertension, Journal of the American College of Cardiology, Pharmacoepidemiology and drug safety, Drug safety, Annals of internal medicine, The Lancet. Digital health, The Lancet. Infectious diseases, The lancet. Diabetes endocrinology, The Lancet. Neurology, The Lancet. Oncology, The Lancet. Respiratory medicine, The lancet. Psychiatry, and The Lancet. Global health*. The full search parameters used are given below. Note that this literature review was performed in 2023 and, as such, the query below may yield additional results if submitted today.

Authors JB and PBR divided the set of returned publications and reviewed the full text of each to determine whether an NU design was in fact employed. For publications employing an NU design, all target-comparator pairs for which results were available were recorded verbatim, and the reviewer attempted to manually adjudicate each to an RxNorm ingredient or ATC level 4 class. In order to accommodate class-based cohorts found in the literature, we additionally generated similarity data for all ATC level 4 classes in the same manner as described for RxNorm ingredients in the main text. In some cases, target or comparator groups were defined as composites of individual ingredients and/or classes, in which case the reviewer created separate entries for each component and noted the mapping as "partial". Cohort similarity scores for all mapped target-comparator pairs were then obtained from the analytic pipeline described in the main text and are available as a separate Microsoft Excel workbook.

### PubMed Query

("marketscan" OR "truven" OR "optum" OR "clinformatics" OR "iqvia" OR "pharmetrics" OR "administrative claims" OR "insurance claims" OR "healthcore" OR "aetna" OR "cigna" OR "humana" OR "sentinel")

AND

("cohort" AND ("comparator" OR "compared" OR "comparative" OR "propensity score" OR "versus"))

AND

("risk" OR "effect" OR "causal" OR "hazard" OR "exposure" OR "RR" OR "HR" OR "IRR" OR "rate ratio")

AND

("2021/01/01"[Date - Publication] : "3000"[Date - Publication])

AND

("Science"[Journal] OR "Nature"[Journal] OR "Nature Medicine"[Journal] OR "The New England journal of medicine"[Journal] OR "JAMA"[Journal] OR "JAMA internal medicine"[Journal] OR "JAMA Surgery"[Journal] OR "JAMA Psychiatry"[Journal] OR "JAMA Cardiology"[Journal] OR "JAMA Pediatrics"[journal] OR "JAMA Neurology"[journal] OR "Lancet"[Journal] OR "BMJ"[Journal] OR "Diabetes care"[Journal] OR "Hypertension"[Journal] OR "Journal of the American College of Cardiology"[Journal] OR "Pharmacoepidemiology and drug safety"[Journal] OR "Drug safety"[Journal] OR "Annals of internal medicine"[Journal] OR "The Lancet. Digital health"[Journal] OR "The Lancet. Infectious diseases"[Journal] OR "The lancet. Diabetes endocrinology"[Journal] OR "The Lancet. Neurology"[Journal] OR "The Lancet. Oncology"[Journal] OR "The Lancet. Respiratory medicine"[Journal] OR "The lancet. Psychiatry"[Journal] OR "The Lancet. Global health"[Journal] )

AND

("Chemicals and Drugs Category"[Mesh]) NOT ("cross sectional") NOT ("Randomized Controlled Trial"[Publication Type] OR "Meta-Analysis"[Publication Type])

### References Returned

17. Faquetti ML, la Torre AM-D, Burkard T, Obozinski G, Burden AM. Identification of polypharmacy

patterns in new-users of metformin using the Apriori algorithm: A novel framework for investigating concomitant drug utilization through association rule mining. Pharmacoepidemiol Drug Saf. 2023;32:366–81.
